# Prevalence trends of diabetes subgroups in US: A data-driven analysis spanning three decades from NHANES (1988-2018)

**DOI:** 10.1101/2020.12.10.20247452

**Authors:** Neftali Eduardo Antonio-Villa, Luisa Fernández-Chirino, Arsenio Vargas-Vázquez, Carlos A. Fermín-Martinez, Carlos A. Aguilar-Salinas, Omar Yaxmehen Bello-Chavolla

## Abstract

**AIMS:** Data-driven diabetes subgroups were proposed as an alternative to address diabetes heterogeneity. However, changes in trends for these subgroups have not been reported. Here, we analyzed trends of diabetes subgroups, stratified by sex, race, education level, age categories and time since diabetes diagnosis in the U.S.

**METHODS:** We used data from consecutive NHANES cycles spanning the 1988-2018 period. Diabetes subgroups (mild obesity-related [MOD], severe-insulin deficient [SIDD], severe-insulin resistant [SIRD], and mild age-related diabetes [MARD]) were classified using validated self-normalizing neural networks. Severe autoimmune-diabetes (SAID) was assessed for NHANES-III. Prevalence was estimated using examination sample weights considering bi-cyclic changes (BC) to evaluate trends and changes over time.

**RESULTS:** Diabetes prevalence in the US increased from 7.5% (95%CI 7.1-7.9) in 1988-1989 to 13.9% (95%CI 13.4-14.4) in 2016-2018 (BC 1.09%, 95%CI 0.98-1.31, p<0.001). Non-Hispanic Blacks had the highest prevalence. Overall, MOD, MARD, and SIDD had an increase during the studied period. Particularly, Non-Hispanic Blacks had sharp increases in MARD and SIDD, Mexican Americans in SIDD, and non-Hispanic Whites in MARD. Males, subjects with secondary/high school, and adults aged 40-64 years had the highest increase in MOD prevalence. Trends in diabetes subgroups sustained after stratifying time since diabetes diagnosis.

**CONCLUSIONS:** Prevalence of diabetes and its subgroups in the U.S. have increased from 1988-2018. These trends were different across sex, ethnicities, education, and age categories, indicating significant heterogeneity in diabetes within the U.S. Obesity burden, population aging, socioeconomic disparities, and lifestyle aspects could be implicated in the uprising trends of diabetes in the U.S.

## INTRODUCTION

Diabetes is a highly heterogeneous disease with complex and various underlying etiologies, which result in diverse clinical courses, treatment response, and risk profiles for chronic micro and macrovascular complications (1). Recent studies have reported steady increases in diabetes prevalence worldwide due to various social, genetic, and lifestyle-related factors (2). As an approach to address diabetes heterogeneity, Ahlqvist et al. proposed the assessment of five data-driven diabetes subgroups, including severe-autoimmune (SAID), mild age-related (MARD), mild obesity-related (MOD), severe-insulin deficient (SIDD) and severe-insulin resistant diabetes (SIRD) (3,4). These data-driven diabetes subgroups have been reproduced independently in other cohorts and have shown unique genetic, risk and clinical profiles which are likely to influence assessments of diabetes heterogeneity and clinical course differently across populations (5).

Although the application of diabetes subgroups within clinical practice could help identify the risk for associated complications and expected positive treatment response as an approach to further personalized medicine in diabetes, its determinants at a population level and its potential usefulness to inform public health policies remain unclear. Previous reports of data-driven diabetes subgroups in the US have largely focused on point-prevalence estimations and implications of diabetes subgroups for mortality (6). However, the implications of these subgroups to understand diabetes heterogeneity and the evolution of diabetes trends according to known modifiers of diabetes have not previously been explored. Here, we analyzed trends of overall diabetes and its proposed data-driven subgroups using the National Health and Nutrition Examination Surveys spanning a three-decade period in the U.S. from 1988 to 2018. We aim to evaluate the epidemiological impact of sociodemographic factors, population aging, and obesity on diabetes and its subgroups stratifying their estimated weighted prevalence by age, sex, ethnicity, education level, time since diabetes diagnosis and body-mass index (BMI) categories.

## METHODS

### Study subjects

We used U.S. citizen data from the 1988-2018 cycles of NHANES. NHANES had the approval of the NCHS Ethics Review Board. Details of response rate, medical questionnaires, anthropometrical and biochemical measurements for each cycle are reported elsewhere (7). Diabetes was defined as glycated hemoglobin (HbA1c) ≥6.5%, fasting plasma glucose ≥7.0 mmol/L (≥126 mg/dL), or previous diabetes diagnosis following current ADA guidelines. Recently diagnosed diabetes was defined as cases with <5 years since diagnosis (8). We analyzed information of adults ≥18 years with complete data to estimate diabetes subgroups. A flowchart of data extraction is available in **Supplementary Figure 1**.

### Diabetes subgroup estimation in NHANES

Prediction of data-driven diabetes subgroups was based on the algorithms previously developed by our study group (9). Briefly, these estimations of diabetes subgroup classification are based on a supervised machine learning algorithm which uses self-normalizing artificial neural networks (SNNN) trained with surrogate metabolic measures to estimate insulin action from population-based studies. Furthermore, the SNNN approach has the advantage of overcoming the limited availability of C-peptide measurements, which are only available for NHANES-III (1988-1994) and NHANES 1999-2006, by implementing insulin measurements to evaluate insulin sensitivity and β-cell function, available for the remaining NHANES 2007-2018 cycles. SNNN models were originally trained using NHANES-III data and were compared to k-means clustering results using the original variables proposed by Ahlqvist et al, thus leading to consistent applications in this dataset (9).

- SNNN-Model 1 was developed considering the original clustering variables including age at diabetes diagnosis, BMI, HbA1c and Homeostasis Model Assessment 2 (HOMA2) metrics calculated using glucose and C-peptide measures to obtain HOMA2-IR and HOMA2-β; the performance metrics (area under the receiving operating characteristic curve, AUROC) of model 1 in NHANES 1999-2006 for all subgroups ranged from AUROC=0.99 to 1.0 when assessed using confusion matrices after SNNN classification compared to k-means clustering with the original variables.
- SNNN-Model 2 uses similar variables as SNNN-Model 1 but estimates HOMA2 metrics using fasting glucose and insulin measurements instead of C-peptide. These models showed adequate performance in NHANES 1999-2006 with AUROC ranging from 0.86 to 0.97 for all diabetes subgroups compared to k-means clustering.

For this work, we used SNNN-Model 1 for the prediction of diabetes subgroups in subjects living with diabetes who had available C-peptide measurements (NHANES-III and NHANES 1999-2006). The SNNN-Model 2 was applied in subjects living with diabetes and available fasting insulin for the remaining NHANES cycles (2007-2018). For the subgroup estimation, we used an electronic-based application previously developed by our group, available at: https://uiem.shinyapps.io/diabetes_clusters_app/. The algorithm does not explicitly consider SAID, as it was conceived as a diagnosis which requires positive anti-glutamic acid decarboxylase antibodies (anti-GAD65). We defined SAID using anti-GAD65 index values >0.069, which were only available for the 1988-1991 NHANES cycles (NHANES-III). Therefore, SAID analyses were only included as a weighted sub-analysis for these cycles.

### Weighted prevalence estimation of diabetes subgroups

Prevalence of diabetes was estimated using examination sample weights from NHANES for participants with available insulin measures; all estimations were conducted using the *survey* R package. To estimate changes across two combined NHANES periods of 4 years (bi-cyclic changes [BC]), we fitted linear regression models using BC as continuous variables to evaluate trends over time and increase statistical power (10). We further performed weighted subgroup analyses for trends stratified by age, sex, ethnicity (codified as Mexican Americans, Non-Hispanic Whites and Non-Hispanic Blacks), education level (codified as primary school/no-education, secondary/high-school/A.A. degree and college/higher), time since diabetes diagnosis (codified as recently diagnosed and long-standing diabetes) and BMI category (codified as normal weight, overweight and obesity).

### Influence of modifying variables on diabetes subgroup trends

To evaluate the effect of sociodemographic factors, population aging, and obesity as categorical modifiers of overall diabetes and its subgroups over time, we implemented mixed-effects linear models including the interaction between the modifier and diabetes subgroup as a fixed effect and a by-cyclic period by diabetes subgroup interaction as a random intercept to consider the dependence of prevalence measures at any given cycle compared to previous years. These models were implemented with the *lme4* R package and p-values were estimated using the Satterthwaite’s degrees of freedom method with the *lmerTest* R package. Statistical significance was established at a p<0.05 threshold. All statistical analyses were conducted using R software version 4.0.3.

## RESULTS

### Study population across NHANES cycles

We extracted complete clinical and biochemical information from 5,489 patients living with diabetes, in which we perform subgroup classification using SNNN. We observed a mean age of 61.2 (±14.3) years, and a male (51.2%), non-Hispanic white (44.3%) and secondary, high-school or AA Degree (41.3%) education predominance. We observed an increase in age and BMI, whilst the proportion of recently diagnosed diabetes, HOMA2-IR and HOMA2- β values decreased across the studied period. Complete sociodemographic, clinical, and biochemical characteristics of patients living with diabetes across NHANES bi-cyclic periods from 1988 to 2018 are presented in **Supplementary Table 1**. Using the SNNN algorithm to classify diabetes subgroups, we identified 1,654 cases (30.1%) categorized as MOD, 1,403 (25.6%) as MARD, 1,290 (23.5%) as SIDD, and 1,142 (20.8%) as SIRD (**Table 1**).

**Table 1:** Sociodemographic, clinical and biochemical characteristics of patients living with diabetes stratified by diabetes subgroups classified from NHAHES cycles from 1988-2018.

| Parameter | All Population<br>(n=5,489) | Diabetes Subgroups |  |  |  | p-value |
| --- | --- | --- | --- | --- | --- | --- |
|  |  | MOD<br>(n=1,654) | MARD<br>(n=1,403) | SIDD<br>(n=1,290) | SIRD<br>(n=1,142) |  |
| Female (%) | 2680 (48.82) | 897 (54.23) | 609 (43.41) | 605 (46.9) | 569 (49.82) | 0.0002 |
| Age (Years) | 61.2 ($\pm 14.3$) | 50.63 ($\pm 13.48$) | 71.34 ($\pm 8.64$) | 58.17 ($\pm 13.14$) | 67.23 ($\pm 9.86$) | <0.001 |
| Mexican American (%) | 1637 (29.82) | 531 (32.10) | 322 (22.95) | 486 (36.67) | 298 (26.09) | <0.001 |
| Non-Hispanic White (%) | 2430 (44.27) | 656 (39.66) | 790 (56.3) | 453 (35.11) | 531 (46.69) | <0.001 |
| Non-Hispanic Black (%) | 1422 (25.91) | 467 (28.23) | 291 (20.74) | 351 (27.21) | 313 (27.41) | <0.001 |
| Primary School or No-Education (%) | 1415 (25.78) | 327 (19.77) | 366 (26.09) | 394 (30.54) | 328 (28.72) | <0.001 |
| Secondary, High-School or AA Degree (%) | 2262 (41.21) | 719 (43.47) | 561 (39.99) | 510 (39.53) | 472 (41.33) | 0.1188 |
| College or Higher (%) | 1812 (33.01) | 608 (36.76) | 476 (33.93) | 386 (29.92) | 342 (29.95) | <0.001 |
| Glycated Hemoglobin (%) | 7.33 ($\pm 1.82$) | 6.62 ($\pm 0.94$) | 6.56 ($\pm 0.81$) | 9.9 ($\pm 1.7$) | 6.41 ($\pm 0.85$) | <0.001 |
| Glucose (mg/dl) | 159.09 ($\pm 65.22$) | 137.74 ($\pm 36.1$) | 138.3 ($\pm 30.58$) | 243.5 ($\pm 71.85$) | 120.19 ($\pm 28.76$) | <0.001 |
| Insulin (mUI) | 23.26 ($\pm 44.17$) | 22.75 ($\pm 24.78$) | 13.3 ($\pm 63.44$) | 26.84 ($\pm 46.15$) | 32.21 ($\pm 30.43$) | <0.001 |
| Weight (Kg) | 86.71 ($\pm 22.36$) | 95.66 ($\pm 25.04$) | 72.68 ($\pm 13.59$) | 86.79 ($\pm 20.85$) | 90.89 ($\pm 20.04$) | <0.001 |
| Height (cm) | 165.97 ($\pm 10.18$) | 166.59 ($\pm 10.23$) | 165.11 ($\pm 10.08$) | 166.56 ($\pm 9.96$) | 165.47 ($\pm 10.36$) | 0.122 |
| Body Mass Index (kg/m<sup>2</sup>) | 31.36 ($\pm 7.09$) | 34.38 ($\pm 8.08$) | 26.56 ($\pm 3.72$) | 31.18 ($\pm 6.4$) | 33.1 ($\pm 6.24$) | <0.001 |
| HOMA2-B | 79.94 ($\pm 110.31$) | 82.49 ($\pm 46.13$) | 51.88 ($\pm 21.71$) | 36.77 ($\pm 36.7$) | 159.5 ($\pm 209.58$) | <0.001 |
| HOMA2-IR | 2.98 ($\pm 4.26$) | 2.64 ($\pm 2.08$) | 1.48 ($\pm 0.93$) | 4.16 ($\pm 7.63$) | 4 ($\pm 2.92$) | <0.001 |
| Time Since Diabetes Diagnosis (Years) | 12.03 ($\pm 11.6$) | 15.46 ($\pm 13.78$) | 8.43 ($\pm 6.94$) | 12.56 ($\pm 9.51$) | 8.07 ($\pm 8.04$) | <0.001 |
| Recently Diagnosed Diabetes (%) | 1028 (18.72) | 898 (54.29) | 856 (61.01) | 498 (38.6) | 797 (69.79) | <0.001 |

*Categorical data is expressed in frequency and absolute proportion. Continuous data is expressed in median (standard deviation) or median (interquartile range) wherever appropriated. \*Percentage is expressed according to available information for diabetes subgroups. Abbreviations: MARD: Mild Age-Related. MOD: Mild-Obesity Related. SIDD: Severe-Insulin Deficient. SIRD: Severe-Insulin Resistant.*

### Trends in diabetes subgroups

In the weighted prevalence analysis, we observed that diabetes has steadily increased in the US from 7.5% (95%CI 7.1-7.9) in 1988-1989 to 13.9% (95%CI 13.4-14.4) in 2016-2018 (BC 1.09%, 95%CI 0.98-1.31, p<0.001). Regarding diabetes subgroups, MOD had the greatest increase in prevalence over time, followed by MARD and SIDD. We observed that SIRD had non-significant increases in prevalence during the three decades period (**Figure 1**). SAID remained stable during the 6 years covered by NHANES-III period in which its prevalence could be assessed (**Supplementary Figure 2**). Distribution of classification variables for data-driven diabetes subgroups are presented in detail within **Supplementary Figure 3**.

**Figure 1:**
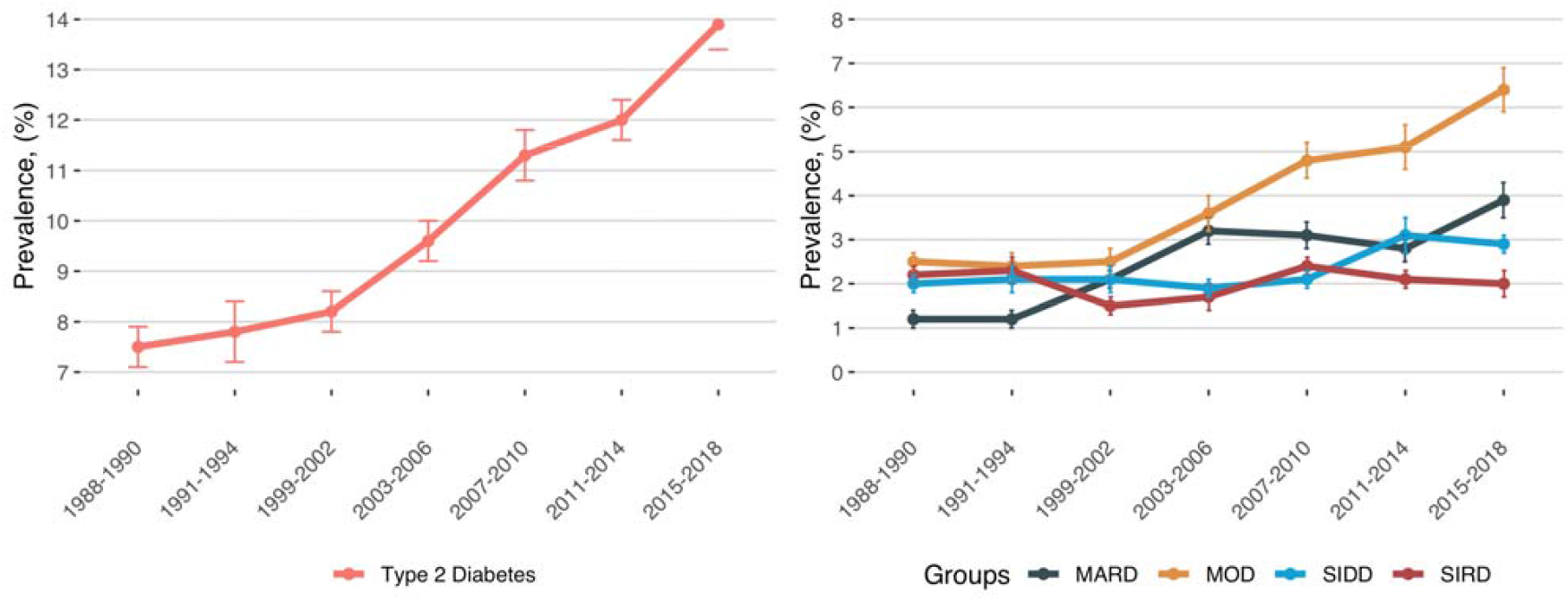
Trends of prevalence for diabetes and its subgroups in the National Health and Nutrition Examination Survey from 1988-2018. Prevalence estimations are weighed according to survey design. *Abbreviations*: MARD, mild age-related diabetes; MOD, mild obesity-related diabetes; SIDD, severe insulin-deficient diabetes; SIRD, severe insulin-resistant diabetes.

### Sex-based trends in diabetes subgroups

We observed the largest increase and proportion in recent years for diabetes prevalence in men compared to women. When evaluating changes in diabetes subgroups, MOD and MARD had an overall increase across sexes, nevertheless only men had increases in SIDD prevalence across time compared to women. SIRD remained unchanged in proportion and prevalence for both sexes (**Supplementary Table 2**; **Figure 2**).

**Figure 2:**
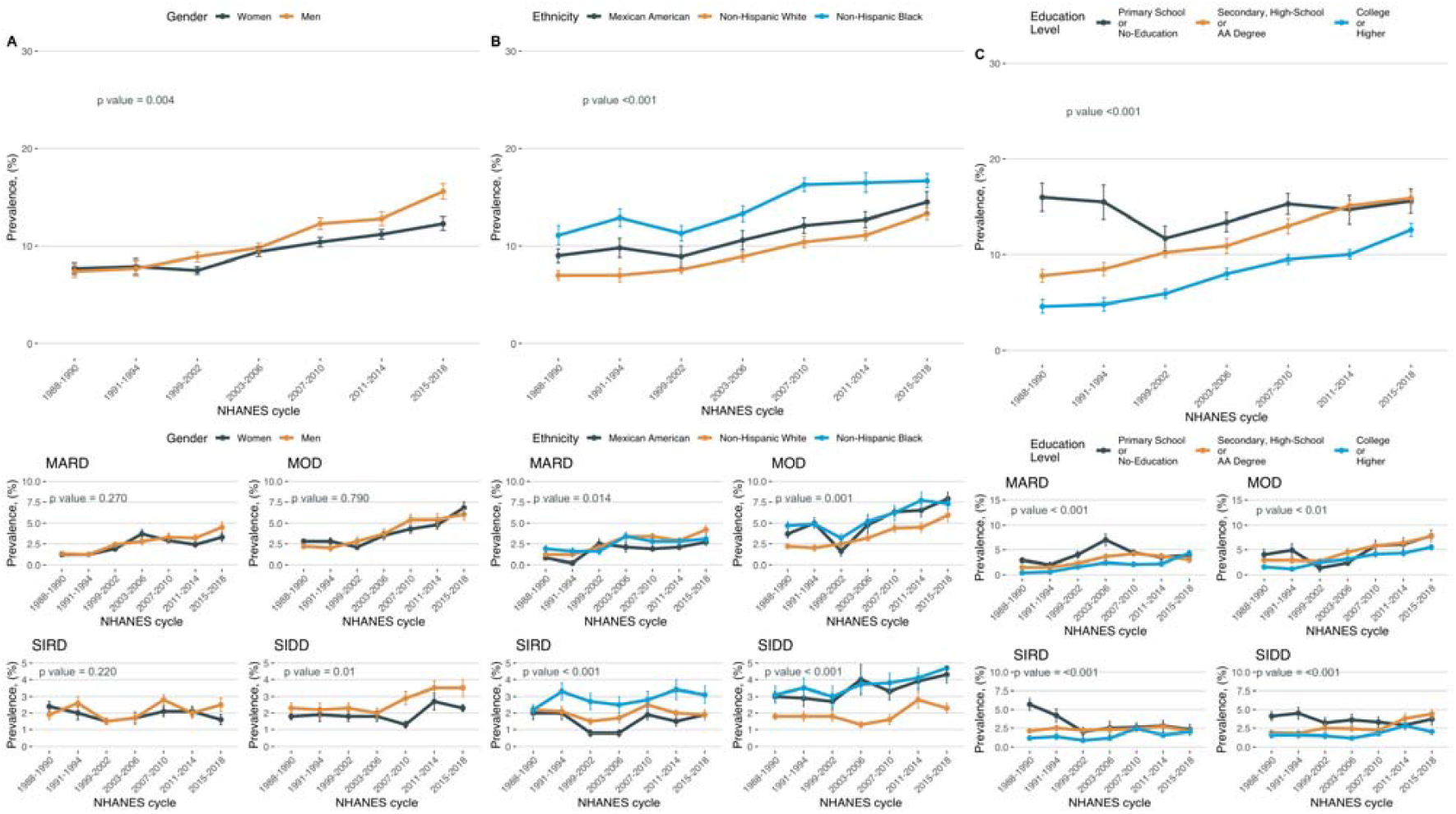
Trends of prevalence for diabetes and its subgroups stratified by ethnicity (A) codified as Mexican Americans, Non-Hispanic Whites and Non-Hispanic Blacks; education level (B) codified as primary school/no-education, secondary/high-school/A.A. degree and college/higher, and age categories (C) in the National Health and Nutrition Examination Survey from 1988-2018. Prevalence estimations are weighed according to survey design. *Abbreviations*: MARD, mild age-related diabetes; MOD, mild obesity-related diabetes; SIDD, severe insulin-deficient diabetes; SIRD, severe insulin-resistant diabetes.

### Diabetes subgroup prevalence trends are different across ethnicities

Overall, ethnicity had meaningful effects on the trajectory of diabetes prevalence trends. Non-Hispanic Blacks and Mexican Americans had the highest prevalence of diabetes compared with Non-Hispanic Whites. Regarding data-driven diabetes subgroups, Non-Hispanic Blacks had marked increased prevalence in MOD, SIDD and SIRD, Mexican Americans in MOD and SIDD and Non-Hispanic Whites in MOD and MARD (**Figure 2; Supplementary Table 3**).

### Lower-educational level impacts on diabetes subgroup trends

We observed significant differences in diabetes prevalence trends when stratifying by education level. Although adults with primary-school/no-education had the highest diabetes prevalence compared with higher education attainment, only subjects with secondary/high-school/A.A. and college or higher education had an increase in diabetes prevalence across time. When assessing weighted prevalence of diabetes subgroups, we observed subjects with college or higher education had an increased in MOD and MARD. In subjects with secondary/high-school/A.A. and college, MOD and SIDD had marked increase in prevalence and finally, in subjects with primary-school/no-education, only MOD shown an increased trend across time. SIRD remained unchanged regardless education attainment (**Figure 2; Supplementary Table 3**).

### Effect of age on diabetes subgroup prevalence trends is modified by ethnicity

Aging has a direct impact on diabetes prevalence. Adults aged >65 years had the highest proportion and increase in diabetes prevalence across our studied period. Assessing diabetes subgroups trends, we observed that adults aged 40-64 had the largest increased in MOD prevalence, while older adults (>65 years) displayed a large increase in MARD and MOD phenotypes over time. Adults aged 18-39 had unchanged prevalence of diabetes or their subgroups (**Supplementary Figure 4 and Supplementary Table 4**).

### Impact of BMI on diabetes subgroup prevalence trends

Adults with obesity had the highest increase in diabetes prevalence compared with overweight and normal-weight subjects. We observed that for normal-weight subjects, SIDD followed an upward trend in prevalence. For adults with overweight and obesity, these upwards trends increased for MOD. Notably, MARD prevalence increased across all BMI categories while SIRD decreased amongst the groups with obesity and overweight (**Supplementary Figure 5 and Supplementary Table 5**).

### Time since diabetes onset modified trends across diabetes subgroups

Finally, we sought to evaluate the effect of recently diagnosed diabetes as a modifier of the overall prevalence and its subgroups. We observed that diabetes with ≥5 years since diagnosis had a sharp increase across our study period, surpassing adults with recently diagnosed diabetes within the last decade, likely as the number of diagnosed individuals also experienced longer lifespans. Furthermore, subjects with recently diagnosed diabetes tend to have the highest proportion and weighted prevalence of MARD, MOD, and SIRD, while subjects that have been living with diabetes for ≥5 years has higher rates of SIDD. We observed that for both categories, MARD and MOD had increases in prevalence over time regardless the time since diabetes diagnosis. SIRD had a decrease in recently diagnosed cases, while SIDD prevalence increased over time in cases with ≥5 years since diagnosis, eventually surpassing cases with recently diagnosed diabetes in the last decade (**Supplementary Figure 6**).

## DISCUSSION

Here, we show an increase in the overall prevalence of diabetes and its data-driven subgroups using U.S. citizen data from consecutive NHANES cycles spanning a three-decade period from 1988 to 2018. Data-driven diabetes subgroup trends were different when considering sex, ethnicity, education level, age, BMI groups and time since diabetes diagnosis, establishing that these factors could have likely influenced the heterogeneity in diabetes profiles across time. Furthermore, our results also support an unequal distribution of the increased burden of this pathology over time on ethnically underrepresented minorities and subjects with lower educational attainments. Overall, our results support the fundamental contribution of socioeconomic factors, obesity, and population aging as major contributors for the overall increase in diabetes prevalence heterogeneity across time.

Our results are supported by the wide epidemiological based studies that have unveiled the contribution of biological, ethnic and social factors associated with diabetes. As previously reported, we observed that men had sharper increases in diabetes prevalence, particularly for SIDD, which could be attributed to sex-based differences in treatment adherence and glycemic control (11). Moreover, we demonstrated that diabetes prevalence varies significantly by ethnicity. Notably, Non-Hispanic Blacks and Mexican Americans had marked increases in MOD, MARD, and SIDD prevalence, while non-Hispanic Whites had increases predominantly on MOD and MARD, reflecting disparities underlying cardio-metabolic burden and glycemic control influenced by ethnicity (12). Our results are consistent with the reported rise in obesity prevalence in the U.S., particularly in Mexican-Americans and non-Hispanic Blacks (13). Previous research has shown that non-Hispanic Blacks and Mexican-Americans develop diabetes at lower BMI and have on average, an earlier age of diabetes onset compared to non-Hispanic Whites, as observed in MOD (14). A previous diabetes subgroup study in Mexicans identified a higher incidence of SIRD and MOD in cases with higher rates of metabolic syndrome traits and a lower incidence of SIDD and MARD, which has reported a higher prevalence in Europeans (9,14). These differences are likely attributable to ethnic-specific genetic differences in metabolic function, differences in age structures across different populations, and ethnic disparities in management and occurrence of cardiometabolic risk factors (15,16). Our study provides relevant insights on ethnic-specific differences in diabetes heterogeneity within the U.S., consistent with previous studies using NHANES-III data, and identifies ethnic groups that may be particularly vulnerable to the rise in obesity and population aging (13). Furthermore, our study shows that education level plays a significant role in modifying diabetes prevalence and heterogeneity over time; most notably, this became apparent for non-Hispanic Whites. The role of education level and ethnicity in modifying the clinical course of diabetes had been previously explored, which could partially explain the differences observed in our study (17). Further pathophysiological, genetic, and socioeconomic determinants in diabetes phenotypes across ethnic groups in the U.S. and in other populations, along with the use of the web-based application to classify patients living with diabetes within its subgroups in real clinical practice scenarios should be assessed to understand their role in shaping diabetes subgroup incidence and long-term glycemic control.

Our study had some strengths and limitations. We used a robust dataset spanning 30 years of NHANES grouping bi-cyclic changes in diabetes prevalence to increase statistical power and evaluate the influence of known modifiers of diabetes prevalence and control. Moreover, we observed that the distribution of diabetes subgroups clustering variables used in our model algorithm followed the expected pattern observed for previous data-driven classifications of diabetes subgroups, confirming the reliability of the SNNN methodology (18,19). Nevertheless, some limitations to be acknowledged include that risk factors related to subgroup incidence cannot be evaluated in a cross-sectional survey, opening further study opportunities. Similarly, we only assessed SAID prevalence in the 1988 to 1994 cycles due to lack of auto-antibody measurements for other NHANES cycles, limiting our ability to study changes in the prevalence of autoimmune diabetes over time. We also acknowledge that criteria for the definition of diabetes have varied over time within the study period, which could have influenced the estimation of diabetes prevalence, particularly for earlier NHANES cycles. Finally, since data required for the identification of data-driven diabetes subgroups are only available for a subset of NHANES participants, we could not explore all possible category combinations to assess the combined effect of all evaluated covariates, which remains an area of opportunity for further assessment.

In summary, the prevalence of diabetes and its subgroups have increased from 1988-2018 amongst all evaluated groups. Obesity is an essential determinant of the increases in diabetes prevalence over time (20); however, our results suggest that this is true primarily for MOD and SIRD, but not MARD nor SIDD. The crescent increase in MOD phenotype should encourage to implement cost-effective strategies to diminish the effect attributable to obesity in the growth of diabetes over time. Along with a rise in diabetes prevalence trends, our results also reflect increases in diabetes heterogeneity, which are likely to complicate treatment standardization and suggest differential effects of public health measures to reduce diabetes burden, which should consider some of the modifiers we explored in this study. This expansion in diabetes heterogeneity and subgroup prevalence over time is likely attributable to multifactorial causes, including but not restricted to increasing obesity rates, population aging, socioeconomic disparities, and lifestyle, which should be assessed longitudinally to inform public policy (21).

## Data Availability

All data sources and R code are available for reproducibility of results at: https://github.com/oyaxbell/clusters_nhanes

## ACKNOWLEDGMENTS

NEAV and AVV are enrolled at the PECEM program of the Faculty of Medicine at UNAM and are supported by CONACyT.

## CONFLICT OF INTERESTS

The authors declare that they have no conflict of interests.

## AUTHORS’ CONTRIBUTIONS

Research idea and study design NEAV, LFC, OYBC, AVV; data acquisition: NEAV, LFC; data analysis/interpretation: NEAV, LFC, OYBC, CAFM, AVV, CAAS; statistical analysis: NEAV, LFC, OYBC; manuscript drafting: NEAV, LFC, OYBC, AVV, CAFM, CAAS; supervision or mentorship: OYBC. Each author contributed important intellectual content during manuscript drafting or revision and accepts accountability for the overall work by ensuring that questions about the accuracy or integrity of any portion of the work are appropriately investigated and resolved.

## Notes

### Competing Interest Statement

The authors have declared no competing interest.

### Funding Statement

This research received no specific funding.

### Author Declarations

NHANES had the approval of the NCHS Ethics Review Board

### Summary of Updates

We reanalyzed the data according to reviewers' comments and included a refined analytic sample

