## Supplementary Material for "Prevalence trends of diabetes subgroups in US: A data-driven analysis spanning three decades from NHANES (1988-2018)"

**SUPPLEMENTARY TABLES**

**Supplementary Table 1:** Sociodemographic, clinical and biochemical characteristics of patients living with diabetes stratified by NHANES bicycles from 1988-2018.

| Parameter |  | NHANES Bicycles | | | | | | | p-value |
| --- | --- | --- | --- | --- | --- | --- | --- | --- | --- |
|  | All population  (n = 5,489) | NHANES-III  1988-1990 (n=920; 16.76%) | NHANES-III  1991-1994 (n=626; 11.4%) | 1999-2002 (n=522; 9.51%) | 2003-2006 (n=592; 10.79%) | 2007-2010 (n=931; 16.96%) | 2011-2014 (n=854; 15.56%) | 2015-2018 (n=1,044; 19.02%) |  |
| Female (%) | 2680 (48.82) | 468 (50.87) | 357 (57.03) | 252 (48.28) | 294 (49.66) | 430 (46.19) | 393 (46.02) | 486 (46.55) | 0.0001 |
| Age (Years) | 61.15 (±14.29) | 60.83 (±15.48) | 59.62 (±15.98) | 61.81 (±13.84) | 62.75 (±14.01) | 61.43 (±13.79) | 60.4 (±13.74) | 61.46 (±13.26) | <0.001 |
| Mexican American (%) | 1637 (29.82) | 275 (29.89) | 188 (30.03) | 174 (33.33) | 156 (26.35) | 305 (32.76) | 209 (24.47) | 330 (31.61) | <0.001 |
| Non-Hispanic White (%) | 2430 (44.27) | 373 (40.54) | 230 (36.74) | 230 (44.06) | 279 (47.13) | 425 (45.65) | 418 (48.95) | 475 (45.5) | <0.001 |
| Non-Hispanic Black (%) | 1422 (25.91) | 272 (29.57) | 208 (33.23) | 118 (22.61) | 157 (26.52) | 201 (21.59) | 227 (26.58) | 239 (22.89) | <0.001 |
| Primary School or No-Education (%) | 1415 (25.78) | 374 (40.65) | 256 (40.89) | 146 (27.97) | 132 (22.3) | 200 (21.48) | 135 (15.81) | 172 (16.48) | <0.001 |
| Secondary, High-School or AA Degree (%) | 2262 (41.21) | 398 (43.26) | 264 (42.17) | 212 (40.61) | 254 (42.91) | 407 (43.72) | 347 (40.63) | 380 (36.4) | <0.001 |
| College or Higher (%) | 1812 (33.01) | 148 (16.09) | 106 (16.93) | 164 (31.42) | 206 (34.8) | 324 (34.8) | 372 (43.56) | 492 (47.13) | <0.001 |
| Glycated Hemoglobin (%) | 7.33 (±1.82) | 7.39 (±1.98) | 7.52 (±1.93) | 7.38 (±1.83) | 7.27 (±1.76) | 7.19 (±1.68) | 7.33 (±1.78) | 7.32 (±1.76) | <0.001 |
| Glucose (mg/dl) | 7.33 (±1.82) | 7.39 (±1.98) | 7.52 (±1.93) | 7.38 (±1.83) | 7.27 (±1.76) | 7.19 (±1.68) | 7.33 (±1.78) | 7.32 (±1.76) | <0.001 |
| Insulin (mUI) | 23.26 (±44.17) | 26.24 (±84.17) | 30.04 (±52.42) | 26.07 (±26.38) | 18.69 (±19.55) | 19.72 (±19.97) | 20.22 (±21.61) | 23.42 (±32.17) | <0.001 |
| Weight (Kg) | 86.71 (±22.36) | 81.46 (±19.75) | 82.41 (±19.25) | 86.75 (±21.92) | 88.93 (±23.03) | 88.3 (±22.5) | 89.32 (±23.46) | 89.07 (±23.88) | <0.001 |
| Height (cm) | 165.97 (±10.18) | 165.49 (±9.88) | 164.17 (±10.21) | 166.39 (±10.11) | 166.68 (±10.39) | 166.41 (±10.61) | 166.47 (±9.98) | 166.06 (±9.98) | <0.001 |
| Body Mass Index (kg/m2) | 31.36 (±7.09) | 29.67 (±6.34) | 30.48 (±6.17) | 31.21 (±6.91) | 31.9 (±7.34) | 31.76 (±7.01) | 32.15 (±7.57) | 32.15 (±7.53) | <0.001 |
| HOMA2-B | 79.94 (±110.31) | 91.63 (±70.86) | 98.6 (±72.44) | 87.07 (±153.59) | 68.3 (±89.66) | 69.83 (±57.42) | 70.86 (±55.18) | 77.95 (±184.65) | <0.001 |
| HOMA2-IR | 2.98 (±4.26) | 3.39 (±2.79) | 3.8 (±8.01) | 3.48 (±5.29) | 2.46 (±3.14) | 2.54 (±3.12) | 2.55 (±2.72) | 2.92 (±3.83) | <0.001 |
| Time Since Diabetes Diagnosis (Years) | 12.03 (±11.6) | 11.21 (±9.89) | 10.3 (±9.94) | 12.22 (±14.06) | 12.86 (±14.05) | 11.84 (±11.59) | 12.04 (±10.89) | 13.12 (±11.18) | <0.001 |
| Recently Diagnosed Diabetes (%) | 1028 (29.64) | 553 (60.11) | 402 (64.22) | 304 (58.24) | 308 (52.03) | 523 (56.18) | 458 (53.63) | 501 (47.99) | <0.001 |
| MARD (%) * | 1403 (25.56) | 173 (18.8) | 93 (14.86) | 178 (34.1) | 207 (34.97) | 251 (26.96) | 224 (26.23) | 277 (26.53) | <0.001 |
| MOD (%) * | 1654 (30.13) | 244 (26.52) | 179 (28.59) | 91 (17.43) | 149 (25.17) | 308 (33.08) | 306 (35.83) | 377 (36.11) | <0.001 |
| SIDD (%) * | 1290 (23.5) | 253 (27.5) | 174 (27.8) | 134 (25.67) | 128 (21.62) | 189 (20.3) | 178 (20.84) | 234 (22.41) | <0.001 |
| SIRD (%) * | 1142 (20.81) | 250 (27.17) | 180 (28.75) | 119 (22.8) | 108 (18.24) | 183 (19.66) | 146 (17.1) | 156 (14.94) | <0.001 |

*Categorical data is expressed in frequency and absolute proportion. Continuous data is expressed in median (standard deviation) or median (interquartile range) wherever appropriated.*

**Percentage is expressed according to available information for diabetes subgroups.*

*Abbreviations*: MARD: Mild Age-Related. MOD: Mild Obesity Related. SIDD: Severe Insulin Deficient. SIRD: Severe Insulin Resistant.

Footnote: *Chi-square test were used to compared categorical data and ANOVA test were used to compare continuous data across NHANES years.*

**Supplementary Table 2:** Estimated prevalence of diabetes and its subgroups stratified sex in NHANES cycles from 1988-1990 to 2015-2018. Trends were estimated using 2-year cycles for bi-cyclic change. *Abbreviations*: MARD: Mild Age-Related. MOD: Mild Obesity Related. SIDD: Severe Insulin Deficient. SIRD: Severe Insulin Resistant. * p value <0.05

| Estimated prevalence (95% CI) | | | | | | | | | | |
| --- | --- | --- | --- | --- | --- | --- | --- | --- | --- | --- |
| Ethnicity | **Men** | | | | | **Women** | | | | |
|  | *Diabetes* | *MARD* | *MOD* | *SIRD* | *SIDD* | *Diabetes* | *MARD* | *MOD* | *SIRD* | *SIDD* |
| 1988-1990 | 7.4  (6.8-8) | 1.2  (1-1.4) | 2.1  (1.8-2.4) | 1.8  (1.5-2.1) | 2.2  (1.9-2.5) | 7.7  (7.1-8.3) | 1.1  (0.9-1.3) | 2.6  (2.3-2.9) | 2.2  (1.9-2.5) | 1.7  (1.5-1.9) |
| 1991-1994 | 7.7  (6.9-8.5) | 1.1  (0.9-1.3) | 2.0  (1.6-2.4) | 2.5  (2.1-2.9) | 2.2  (1.8-2.6) | 7.9  (7.1-8.7) | 1.2  (1-1.4) | 2.7  (2.3-3.1) | 1.9  (1.5-2.3) | 1.9  (1.6-2.2) |
| 1999-2002 | 8.9  (8.4-9.4) | 2.4  (2.1-2.7) | 2.8  (2.3-3.3) | 1.5  (1.3-1.7) | 2.3  (2-2.6) | 7.5  (7.1-7.9) | 1.9  (1.6-2.2) | 2.1  (1.8-2.4) | 1.5  (1.3-1.7) | 1.8  (1.5-2.1) |
| 2003-2006 | 9.8  (9.3-10.3) | 2.8  (2.4-3.2) | 3.7  (3.1-4.3) | 1.7  (1.3-2.1) | 2.0  (1.7-2.3) | 9.4  (8.9-9.9) | 3.7  (3.2-4.2) | 3.5  (3-4) | 1.7  (1.4-2) | 1.8  (1.5-2.1) |
| 2007-2010 | 12.3  (11.7-12.9) | 3.3  (2.8-3.8) | 5.4  (4.8-6) | 2.8  (2.5-3.1) | 2.9  (2.5-3.3) | 10.4  (9.9-10.9) | 2.9  (2.7-3.1) | 4.3  (3.8-4.8) | 2.1  (1.8-2.4) | 1.3  (1.1-1.5) |
| 2011-2014 | 12.8  (12.1-13.5) | 3.2  (2.8-3.6) | 5.4  (4.7-6.1) | 2.0  (1.7-2.3) | 3.5  (3.1-3.9) | 11.2  (10.7-11.7) | 2.4  (2.1-2.7) | 4.8  (4.3-5.3) | 2.1  (1.8-2.4) | 2.7  (2.2-3.2) |
| 2015-2018 | 15.6  (14.8-16.4) | 4.5  (3.9-5.1) | 6.0  (5.4-6.6) | 2.5  (2.1-2.9) | 3.5 (3-4) | 12.3  (11.6-13) | 3.3  (2.8-3.8) | 6.8  (6.1-7.5) | 1.6  (1.3-1.9) | 2.3  (2.1-2.5) |
| Bi-Cyclic change | 1.36  (1.1-1.6) | 0.53  (0.39-0.67) | 0.75  (0.57-0.92) | 0.08  (0.01-0.26) | 0.25  (0.11-0.39) | 0.83  (0.60-1.1) | 0.36  (0.09-0.62) | 0.67  (0.39-0.96) | -0.028  (-0.13-0.08) | 0.10  (-0.13 0.08) |
| p-value | <0.001 | <0.001 | <0.001 | 0.405 | 0.013 | <0.001 | 0.045 | 0.005 | 0.628 | 0.256 |

*Footnote: The prevalence was estimated using the examination sample weights from NHANES for participants with available insulin measures according to: Ingram, D. D. et al (2018). National Center for Health Statistics Guidelines for Analysis of Trends. Vital and health statistics. Series 2, Data evaluation and methods research, (179), 1–71. Bi-cyclic changes were estimated using mixed-effects linear models including the interaction between the modifier and diabetes subgroup as a fixed effect and a by-cyclic period by diabetes subgroup interaction as a random intercept.*

**Supplementary Table 3:** Estimated prevalence of diabetes and its subgroups stratified ethnicity in NHANES cycles from 1988-1990 to 2015-2018. Trends were estimated using 2-year cycles for bi-cycle change. *Abbreviations*: MARD: Mild Age-Related. MOD: Mild Obesity Related. SIDD: Severe Insulin Deficient. SIRD: Severe Insulin Resistant. * p value <0.05

| Ethnicity | Non-Hispanic Blacks | | | | | Non-Hispanic Whites | | | | | Mexican Americans | | | | |
| --- | --- | --- | --- | --- | --- | --- | --- | --- | --- | --- | --- | --- | --- | --- | --- |
|  | *Diabetes* | *MARD* | *MOD* | *SIRD* | *SIDD* | *Diabetes* | *MARD* | *MOD* | *SIRD* | *SIDD* | *Diabetes* | *MARD* | *MOD* | *SIRD* | *SIDD* |
| 1988-1990 | 11.1 (10.1-12.1) | 1.7 (1.3-2.1) | 4.3 (3.8-4.8) | 2 (1.8-2.2) | 2.8 (2.4-3.2) | 7.0 (6.5-7.5) | 1.1 (0.9-1.3) | 2.1 (1.8-2.4) | 2.1 (1.9-2.3) | 1.8 (1.6-2) | 9.0 (8.3-9.7) | 0.8 (0.6-1) | 3.5 (3-4) | 1.9 (1.5-2.3) | 2.9 (2.6-3.2) |
| 1991-1994 | 12.9 (12-13.8) | 1.6 (1.2-2) | 4.6 (4.2-5) | 3.2 (2.7-3.7) | 3.3 (2.8-3.8) | 7 (6.3-7.7) | 1.1 (0.9-1.3) | 1.9 (1.5-2.3) | 2.1 (1.8-2.4) | 1.8 (1.5-2.1) | 9.8 (8.8-10.8) | 0.2 (0.1-0.3) | 4.8 (4.2-5.4) | 2 (1.6-2.4) | 2.8 (2.3-3.3) |
| 1999-2002 | 11.3 (10.5-12.1) | 1.6 (1.2-2) | 3.2 (2.7-3.7) | 2.7 (2.2-3.2) | 3 (2.4-3.6) | 7.6 (7.2-8) | 2.1 (1.9-2.3) | 2.5 (2.1-2.9) | 1.5 (1.3-1.7) | 1.8 (1.5-2.1) | 8.9 (7.8-10) | 2.5 (1.9-3.1) | 1.6 (1-2.2) | 0.8 (0.6-1) | 2.7 (2.2-3.2) |
| 2003-2006 | 13.3 (12.5-14.1) | 3.4 (2.9-3.9) | 5.2 (4.4-6) | 2.5 (2-3) | 3.7 (3.2-4.2) | 8.9 (8.4-9.4) | 3.4 (3-3.8) | 3.2 (2.8-3.6) | 1.7 (1.4-2) | 1.3 (1.1-1.5) | 10.6 (9.6-11.6) | 2.1 (1.5-2.7) | 4.8 (3.8-5.8) | 0.8 (0.6-1) | 4 (3.1-4.9) |
| 2007-2010 | 16.3 (15.6-17) | 2.8 (2.3-3.3) | 6.2 (5.3-7.1) | 2.8 (2.3-3.3) | 3.8 (3.2-4.4) | 10.4 (9.8-11) | 3.4 (3-3.8) | 4.4 (3.9-4.9) | 2.5 (2.3-2.7) | 1.6 (1.3-1.9) | 12.1 (11.3-12.9) | 1.9 (1.6-2.2) | 6.3 (5.8-6.8) | 1.9 (1.6-2.2) | 3.3 (2.8-3.8) |
| 2011-2014 | 16.5 (15.5-17.5) | 2.8 (2.4-3.2) | 7.7 (6.7-8.7) | 3.4 (2.8-4) | 4.1 (3.5-4.7) | 11.1 (10.6-11.6) | 2.9 (2.6-3.2) | 4.5 (3.9-5.1) | 2 (1.8-2.2) | 2.8 (2.3-3.3) | 12.7 (11.9-13.5) | 2.1 (1.8-2.4) | 6.5 (5.7-7.3) | 1.5 (1.2-1.8) | 3.9 (3.4-4.4) |
| 2015-2018 | 16.7 (16-17.4) | 3.1 (2.5-3.7) | 7.3 (6.6-8) | 3.1 (2.6-3.6) | 4.7 (4.2-5.2) | 13.3 (12.7-13.9) | 4.2 (3.7-4.7) | 5.9 (5.2-6.6) | 1.9 (1.6-2.2) | 2.3 (2-2.6) | 14.5 (13.4-15.6) | 2.7 (2.3-3.1) | 7.9 (7.1-8.7) | 1.9 (1.6-2.2) | 4.3 (3.8-4.8) |
| Bi-Cyclic change | 1.0 (0.6-1.4) | 0.3 (0.1-0.4) | 0.58 (0.2-1.0) | 0.10  (-0.01-0.3) | 0.24 (0.1-0.3) | 1.1 (0.8-1.3) | 0.5 (0.3-0.7) | 0.7 (0.5-0.8) | 0.0 (-0.1-0.1) | 0.1 (-0.1-0.3) | 0.91 (0.6-1.2) | 0.3 (0.1-0.6) | 0.7 (0.2-1.3) | 0.01 (-0.2-0.2) | 0.23 (0.1-0.4) |
| p-value | 0.004 | 0.039 | 0.029 | 0.139 | 0.001 | <0.001 | 0.004 | <0.001 | 0.918 | 0.233 | 0.002 | 0.052 | 0.036 | 0.975 | 0.020 |

*Footnote: The prevalence was estimated using the examination sample weights from NHANES for participants with available insulin measures according to: Ingram, D. D. et al (2018). National Center for Health Statistics Guidelines for Analysis of Trends. Vital and health statistics. Series 2, Data evaluation and methods research, (179), 1–71. Bi-cyclic changes were estimated using mixed-effects linear models including the interaction between the modifier and diabetes subgroup as a fixed effect and a by-cyclic period by diabetes subgroup interaction as a random intercept.*

**Supplementary Table 3:** Estimated prevalence of diabetes and its subgroups stratified education level in NHANES cycles from 1988-1990 to 2015-2018. Trends were estimated using 2-year cycles for bi-cycle change. *Abbreviations*: MARD: Mild Age-Related. MOD: Mild Obesity Related. SIDD: Severe Insulin Deficient. SIRD: Severe Insulin Resistant. * p value <0.05

| Estimated prevalence (95% CI) | | | | | | | | | | | | | | | |
| --- | --- | --- | --- | --- | --- | --- | --- | --- | --- | --- | --- | --- | --- | --- | --- |
| Education  categories | Primary School or No-Education | | | | | Secondary, High-School or AA Degree | | | | | College or Higher | | | | |
|  | *Diabetes* | *MARD* | *MOD* | *SIRD* | *SIDD* | *Diabetes* | *MARD* | *MOD* | *SIRD* | *SIDD* | *Diabetes* | *MARD* | *MOD* | *SIRD* | *SIDD* |
| 1988-1990 | 16.0  (14.5-17.5) | 2.9  (2.4-3.4) | 4.0  (2.9-5.1) | 5.7  (4.9-6.5) | 4.1  (3.5-4.7) | 7.8 (7.1-8.5) | 1.5 (1.2-1.8) | 2.9 (2.6-3.2) | 2.1 (1.8-2.4) | 1.9 (1.6-2.2) | 4.6 (3.9-5.3) | 0.4 (0.3-0.5) | 1.6 (1.2-2.0) | 1.2 (0.9-1.5) | 1.6 (1.2-2.0) |
| 1991-1994 | 15.5 (13.7-17.3) | 2.0  (1.5-2.5) | 4.9  (3.6-6.2) | 4.2  (3.3-5.1) | 4.5  (3.7-5.3) | 8.5 (7.8-9.2) | 1.5 (1.2-1.8) | 2.9 (2.3-3.5) | 2.5 (2.1-2.9) | 1.8 (1.4-2.2) | 4.8 (4.1-5.5) | 0.6 (0.4-0.8) | 1.2 (0.9-1.5) | 1.4 (1-1.8) | 1.6 (1.2-2.0) |
| 1999-2002 | 11.7 (10.4-13) | 4.0  (3.3-4.7) | 1.4  (0.6-2.2) | 2.0  (1.4-2.6) | 3.2  (2.5-3.9) | 10.2 (9.7-10.7) | 2.3 (1.9-2.7) | 2.7 (2.2-3.2) | 2.2 (1.9-2.5) | 2.5 (1.9-3.1) | 5.9 (5.4-6.4) | 1.6 (1.4-1.8) | 2.5 (2.1-2.9) | 0.9 (0.7-1.1) | 1.5 (1.1-1.9) |
| 2003-2006 | 13.4 (12.4-14.4) | 7.0  (5.8-8.2) | 2.4  (1.9-2.9) | 2.5  (1.6-3.4) | 3.6  (2.8-4.4) | 10.9 (10.1-11.7) | 3.6 (3-4.2) | 4.5 (3.7-5.3) | 2.3 (1.9-2.7) | 2.4 (2-2.8) | 8.0 (7.4-8.6) | 2.4 (2.1-2.7) | 3.1 (2.7-3.5) | 1.2 (1-1.4) | 1.2 (0.9-1.5) |
| 2007-2010 | 15.3 (14.2-16.4) | 4.4  (3.8-5) | 5.8  (4.7-6.9) | 2.6  (2-3.2) | 3.3  (2.6-4) | 13 (12.2-13.8) | 4.2 (3.7-4.7) | 5.8 (5.3-6.3) | 2.4 (2-2.8) | 2.2 (1.9-2.5) | 9.5 (9-10) | 2.1 (1.8-2.4) | 4.1 (3.5-4.7) | 2.4 (2.1-2.7) | 1.8 (1.5-2.1) |
| 2011-2014 | 14.7 (13.2-16.2) | 3.5  (2.8-4.2) | 6.1  (4.8-7.4) | 2.8  (2.1-3.5) | 2.9  (2.3-3.5) | 15.1 (14.5-15.7) | 3.7 (3.3-4.1) | 6.4 (5.6-7.2) | 2.7 (2.4-3) | 3.8 (3.1-4.5) | 10 (9.5-10.5) | 2.2 (1.8-2.6) | 4.3 (3.7-4.9) | 1.6 (1.3-1.9) | 2.8 (2.4-3.2) |
| 2015-2018 | 15.6 (14.3-16.9) | 3.9  (3.2-4.6) | 7.8  (6.6-9) | 2.3  (1.6-3) | 3.7  (2.9-4.5) | 15.9 (15.2-16.6) | 3.0 (2.5-3.5) | 7.7 (6.6-8.8) | 2.1 (1.7-2.5) | 4.4 (3.8-5) | 12.6 (11.9-13.3) | 4.3 (3.7-4.9) | 5.5 (4.9-6.1) | 2.0 (1.6-2.4) | 2.0 (1.8-2.2) |
| Bi-Cyclic change | 0.03  (-0.59-0.65) | 0.23  (-0.37 to 0.83) | 0.65  (-0.04 to 1.34) | -0.44  (-0.81 to -0.072) | -0.15  (-0.33 to 0.02) | 1.44  (1.26-1.6) | 0.39  (0.10-0.66) | 0.87  (0.63-1.12) | 0.02  (-0.06 to 0.11) | 0.40  (0.20-0.59) | 1.35  (1.12-1.59) | 0.55  (0.33-0.77) | 0.69  (0.54-0.84) | 0.15  (-0.001 to 0.31) | 0.14  (-0.02 to 0.31) |
| p-value | 0.93 | 0.489 | 0.126 | 0.066 | 0.151 | <0.001 | 0.044 | 0.001 | 0.653 | 0.011 | <0.001 | 0.004 | <0.001 | 0.121 | 0.164 |

*Footnote: The prevalence was estimated using the examination sample weights from NHANES for participants with available insulin measures according to: Ingram, D. D. et al (2018). National Center for Health Statistics Guidelines for Analysis of Trends. Vital and health statistics. Series 2, Data evaluation and methods research, (179), 1–71. Bi-cyclic changes were estimated using mixed-effects linear models including the interaction between the modifier and diabetes subgroup as a fixed effect and a by-cyclic period by diabetes subgroup interaction as a random intercept.*

**Supplementary Table 4:** Estimated prevalence of T2D and its subgroups stratified age categories in NHANES cycles from 1999-20021988-1990 to 2015-2018. Trends were estimated using 2-year cycles for Bi-Cyclic change. *Abbreviations*: MARD: Mild Age-Related. MOD: Mild Obesity Related. SIDD: Severe Insulin Deficient. SIRD: Severe Insulin Resistant. * p value <0.05

| Estimated prevalence (95% CI) | | | | | | | | | | | | | | | |
| --- | --- | --- | --- | --- | --- | --- | --- | --- | --- | --- | --- | --- | --- | --- | --- |
| Age categories | **18-39 years** | | | | | **40-64 years** | | | | | **≥65 years** | | | | |
|  | *Diabetes* | *MARD* | *MOD* | *SIRD* | *SIDD* | *Diabetes* | *MARD* | *MOD* | *SIRD* | *SIDD* | *Diabetes* | *MARD* | *MOD* | *SIRD* | *SIDD* |
| 1988-1990 | 2.4 (2-2.8) | - | 1.7 (1.4-2) | - | 0.8 (0.5-1.1) | 10.4 (9.6-11.2) | 0.9 (0.7-1.1) | 3.8 (3.3-4.3) | 2.6 (2.3-2.9) | 2.6 (2.3-2.9) | 17.6 (16.1-19.1) | 5.2 (4.5-5.9) | 1.2 (0.9-1.5) | 6.7 (5.9-7.5) | 3.8 (3.2-4.4) |
| 1991-1994 | 2.5 (2-3) | - | 2 (1.5-2.5) | - | 0.3 (0.1-0.5) | 10.6 (9.6-11.6) | 0.7 (0.5-0.9) | 3.4 (2.8-4) | 3 (2.5-3.5) | 3.1 (2.6-3.6) | 17.6 (16-19.2) | 5.5 (4.7-6.3) | 0.8 (0.5-1.1) | 6.5 (5.6-7.4) | 4.3 (3.6-5) |
| 1999-2002 | 1.6 (1.3-1.9) | - | 1.2 (0.9-1.5) | 0.1 (0-0.2) | 0.8 (0.6-1) | 10.9 (10.2-11.6) | 1.5 (1.1-1.9) | 4.6 (3.9-5.3) | 1.9 (1.6-2.2) | 2.7 (2.3-3.1) | 20.3 (19.5-21.1) | 10.4 (9.3-11.5) | 0 (0-0) | 4.9 (4.2-5.6) | 4.2 (3.2-5.2) |
| 2003-2006 | 1.7 (1.4-2) | - | 1.4 (1.1-1.7) | - | 0.8 (0.6-1) | 11.9 (11.3-12.5) | 2.2 (1.8-2.6) | 6 (5.2-6.8) | 1 (0.7-1.3) | 2.6 (2.2-3) | 23 (21.8-24.2) | 13.7 (12.3-15.1) | 2.1 (1.7-2.5) | 7.2 (5.9-8.5) | 2.5 (2-3) |
| 2007-2010 | 2 (1.8-2.2) | - | 1.8 (1.5-2.1) | - | 0.8 (0.6-1) | 13.5 (12.8-14.2) | 1.3 (0.9-1.7) | 7.8 (6.9-8.7) | 2.4 (2.1-2.7) | 2.8 (2.4-3.2) | 27.5 (26.2-28.8) | 14.5 (13.5-15.5) | 3.5 (3-4) | 7.7 (6.8-8.6) | 3 (2.5-3.5) |
| 2011-2014 | 2.1 (1.8-2.4) | - | 2.4 (2-2.8) | - | 0.6 (0.4-0.8) | 15.2 (14.4-16) | 1.7 (1.4-2) | 7.8 (6.9-8.7) | 2.2 (1.8-2.6) | 4.3 (3.6-5) | 25.4 (24.5-26.3) | 10.9 (9.8-12) | 4.1 (3.4-4.8) | 5.8 (5.1-6.5) | 5.2 (4.3-6.1) |
| 2015-2018 | 2.6 (2.3-2.9) | - | 2.6 (2.1-3.1) | - | 0.7 (0.5-0.9) | 17.1 (16.2-18) | 2.6 (2.1-3.1) | 10.1 (9-11.2) | 2.1 (1.5-2.7) | 4 (3.5-4.5) | 28.6 (27.4-29.8) | 13.3 (12.1-14.5) | 5.4 (4.6-6.2) | 5.4 (4.5-6.3) | 4.3 (3.8-4.8) |
| Bi-Cyclic change | 0.0 (-0.1-0.2) | - | 0.2 (-0.01-0.3) | -0.003 (-0.02-0.01) | 0.01 (-0.1-0.1) | 1.1 (0.8-1.4) | 0.2 (0.1-0.4) | 1.1 (0.8-1.4) | -0.1 (0-0.3-0.1) | 0.2 (0.04-0.43) | 2.0 (1.4-2.6) | 1.4 (0.5-2.3) | 0.8 (0.5-1.2) | -0.1 (-0.5-0.3) | 0.1 (-0.3-0.4) |
| p-value | 0.933 | - | 0.133 | 0.661 | 0.791 | <0.001 | 0.036 | <0.001 | 0.486 | 0.060 | <0.001 | 0.033 | 0.006 | 0.678 | 0.670 |

*Footnote: The prevalence was estimated using the examination sample weights from NHANES for participants with available insulin measures according to: Ingram, D. D. et al (2018). National Center for Health Statistics Guidelines for Analysis of Trends. Vital and health statistics. Series 2, Data evaluation and methods research, (179), 1–71. Bi-cyclic changes were estimated using mixed-effects linear models including the interaction between the modifier and diabetes subgroup as a fixed effect and a by-cyclic period by diabetes subgroup interaction as a random intercept.*

**Supplementary Table 5:** Estimated prevalence of T2D and its subgroups stratified by BMI categories in NHANES cycles from 1988-1990 to 2015-2018. Trends were estimated using 2-year cycles for Bi-Cyclic change. *Abbreviations*: MARD: Mild Age-Related. MOD: Mild Obesity Related. SIDD: Severe Insulin Deficient. SIRD: Severe Insulin Resistant. * p value <0.05

| BMI categories | Normal-Weight | | | | | Overweight | | | | | Obese | | | | |
| --- | --- | --- | --- | --- | --- | --- | --- | --- | --- | --- | --- | --- | --- | --- | --- |
|  | *Diabetes* | *MARD* | *MOD* | *SIRD* | *SIDD* | *Diabetes* | *MARD* | *MOD* | *SIRD* | *SIDD* | *Diabetes* | *MARD* | *MOD* | *SIRD* | *SIDD* |
| 1988-1990 | 3.7 (3.3-4.1) | 1.2 (1-1.4) | 1.3 (1-1.6) | 0.5 (0.4-0.6) | 0.8 (0.6-1) | 7.7 (7.1-8.3) | 1.5 (1.2-1.8) | 2 (1.7-2.3) | 1.8 (1.6-2) | 2.3 (1.8-2.8) | 16.9 (15.8-18) | 0.6 (0.4-0.8) | 5.6 (5-6.2) | 6.1 (5.4-6.8) | 4.2 (3.6-4.8) |
| 1991-1994 | 2.2 (1.9-2.5) | 1.4 (1.1-1.7) | 0.3 (0.2-0.4) | 0.2 (0.1-0.3) | 0.4 (0.3-0.5) | 8.5 (7.5-9.5) | 1.5 (1.2-1.8) | 2.5 (2-3) | 2.4 (1.9-2.9) | 2 (1.6-2.4) | 17.2 (15.7-18.7) | 0.1 (0-0.2) | 6 (4.9-7.1) | 5.5 (4.7-6.3) | 5 (4.1-5.9) |
| 1999-2002 | 3.3 (2.9-3.7) | 2.2 (1.8-2.6) | 0.6 (0.3-0.9) | 0.2 (0.1-0.3) | 0.8 (0.6-1) | 7.4 (6.9-7.9) | 3 (2.7-3.3) | 1.5 (1.1-1.9) | 1.2 (0.9-1.5) | 2 (1.6-2.4) | 14.3 (13.5-15.1) | 1.2 (0.9-1.5) | 6.2 (5.3-7.1) | 3.7 (3.2-4.2) | 3.9 (3.2-4.6) |
| 2003-2006 | 4 (3.5-4.5) | 3.3 (2.7-3.9) | 0.5 (0.3-0.7) | 0.4 (0.2-0.6) | 0.6 (0.5-0.7) | 8.5 (7.9-9.1) | 4.3 (3.7-4.9) | 1.7 (1.3-2.1) | 1.1 (0.8-1.4) | 1.8 (1.4-2.2) | 16.3 (15.5-17.1) | 2.2 (1.8-2.6) | 8.7 (7.7-9.7) | 3.7 (3.2-4.2) | 3.3 (2.8-3.8) |
| 2007-2010 | 4.3 (3.9-4.7) | 3.3 (2.7-3.9) | 1.1 (0.8-1.4) | 0.3 (0.1-0.5) | 0.9 (0.7-1.1) | 8.2 (7.6-8.8) | 4 (3.6-4.4) | 2.4 (2-2.8) | 1.6 (1.2-2) | 1.7 (1.4-2) | 20.6 (19.7-21.5) | 2.1 (1.8-2.4) | 11.1 (9.9-12.3) | 5.4 (4.8-6) | 3.7 (3.2-4.2) |
| 2011-2014 | 4.7 (4.2-5.2) | 2.7 (2.3-3.1) | 1.3 (1-1.6) | 0.1 (0.1-0.1) | 0.9 (0.6-1.2) | 9.6 (9-10.2) | 4.5 (4-5) | 3.4 (2.8-4) | 1.3 (1-1.6) | 2 (1.7-2.3) | 20.2 (19.4-21) | 1.3 (1-1.6) | 10 (8.9-11.1) | 4.5 (3.9-5.1) | 6.1 (5-7.2) |
| 2015-2018 | 5 (4.6-5.4) | 3.6 (3-4.2) | 1 (0.8-1.2) | 0.3 (0.2-0.4) | 1.5 (1.1-1.9) | 11.7 (10.9-12.5) | 5.8 (4.9-6.7) | 3.8 (3.2-4.4) | 1.1 (0.7-1.5) | 2.1 (1.8-2.4) | 21.5 (20.6-22.4) | 2.6 (2.2-3) | 12.6 (11.4-13.8) | 4.1 (3.5-4.7) | 4.6 (4.1-5.1) |
| Bi-Cyclic change | 0.3 (0.1-0.6) | 0.4 (0.2-0.6) | 0.1 (-0.1-0.2) | -0.02 (-0.1-0.02) | 0.1 (0.02-0.2) | 0.5 (0.2-0.9) | 0.7 (0.5-0.9) | 0.3 (0.05-0.5) | -0.1(-0.3-0.01) | -0.03 (-0.1-0.04) | 0.9 (0.2-1.6) | 0.3 (0.1-0.6) | 1.2 (0.8-1.6) | -0.2 (-0.6-0.1) | 0.1 (-0.2-0.5) |
| p-value | 0.026 | 0.009 | 0.504 | 0.372 | 0.066 | 0.033 | <0.001 | 0.062 | 0.128 | 0.434 | 0.047 | 0.034 | 0.001 | 0.244 | 0.568 |

*Footnote: The prevalence was estimated using the examination sample weights from NHANES for participants with available insulin measures according to: Ingram, D. D. et al (2018). National Center for Health Statistics Guidelines for Analysis of Trends. Vital and health statistics. Series 2, Data evaluation and methods research, (179), 1–71. Bi-cyclic changes were estimated using mixed-effects linear models including the interaction between the modifier and diabetes subgroup as a fixed effect and a by-cyclic period by diabetes subgroup interaction as a random intercept.*

**Supplementary Figure 1:** Flow-diagram of adults included from data collected of the National Health and Nutrition Surveys from 1988-2018.

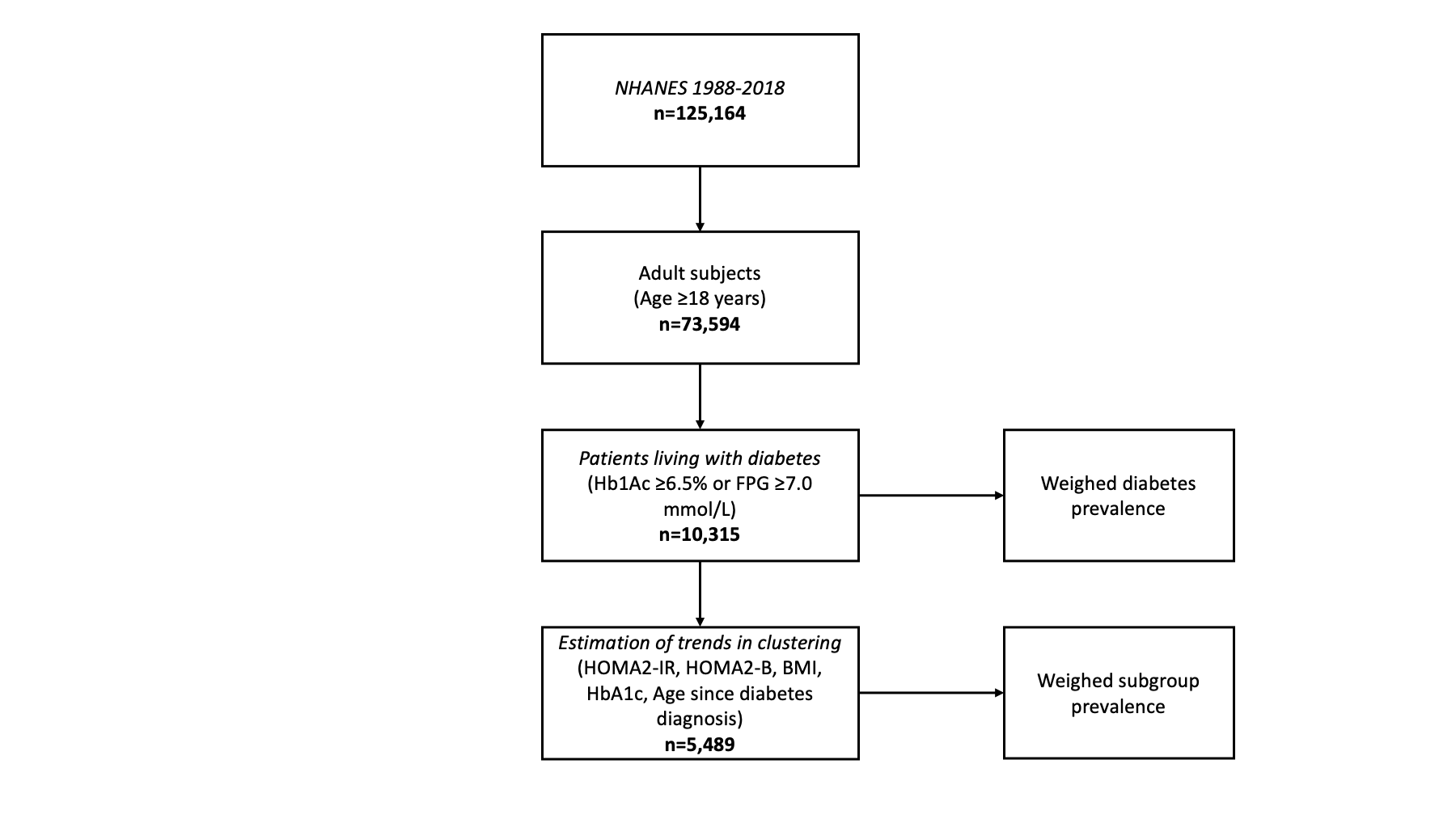

*Abbreviations*: NHANES= National Health and Nutrition Surveys; HbA1c= Glycated hemoglobin; FPG= Fasting plasma glucose; HOMA2-IR= Homeostatic model for Insulin Resistance; HOMA2-B= Homeostatic model for Beta Cell Secretion; BMI= Body Mass Index.

**Supplementary Figure 2:** Weighted prevalence (A) and absolute proportion (B) of diabetes subgroups in the National Health and Nutrition Surveys from 1988-1994 (NHNAES-III). Distribution of diabetes subgroups according to glycated hemoglobin (C), HOMA2-IR (D), HOMA2-B (E), age at diabetes onset (F) and body mass index (G).

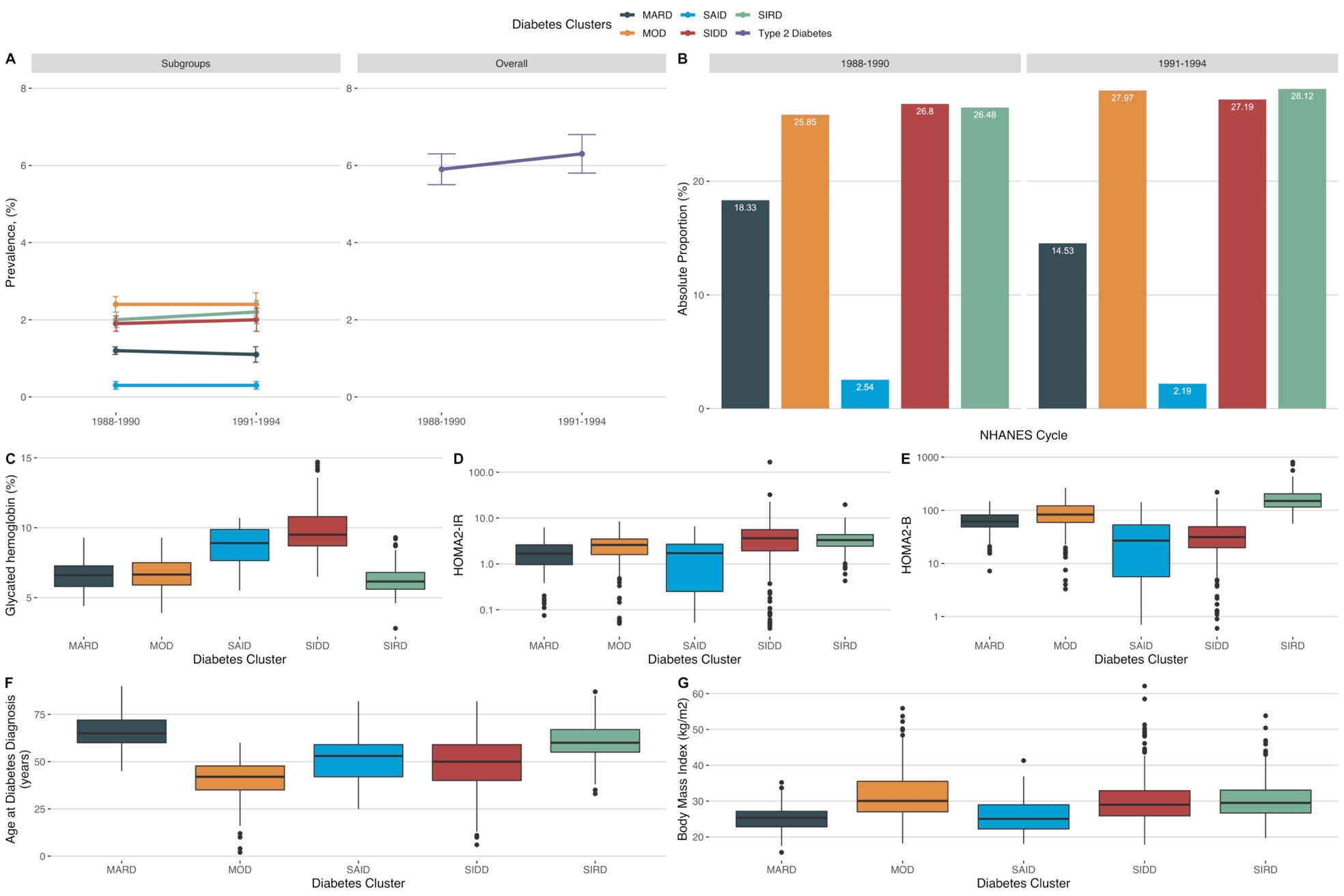

*Abbreviations*: MARD: Mild Age-Related. MOD: Mild Obesity Related. SIDD: Severe Insulin Deficient. SIRD: Severe Insulin Resistant

**Supplementary Figure 3:** Proportion of diabetes subgroups in the National Health and Nutrition Surveys from 1988-2018 (A). Diabetes groups characterization. Distribution of diabetes subgroups according to glycated hemoglobin (B), HOMA2-IR (C), HOMA2-B (D), age at diabetes onset (E) and body mass index (F) in the National Health and Nutrition Surveys from 1999-2018. *Abbreviations*: MARD: Mild Age-Related. MOD: Mild Obesity Related. SIDD: Severe Insulin Deficient. SIRD: Severe Insulin Resistant

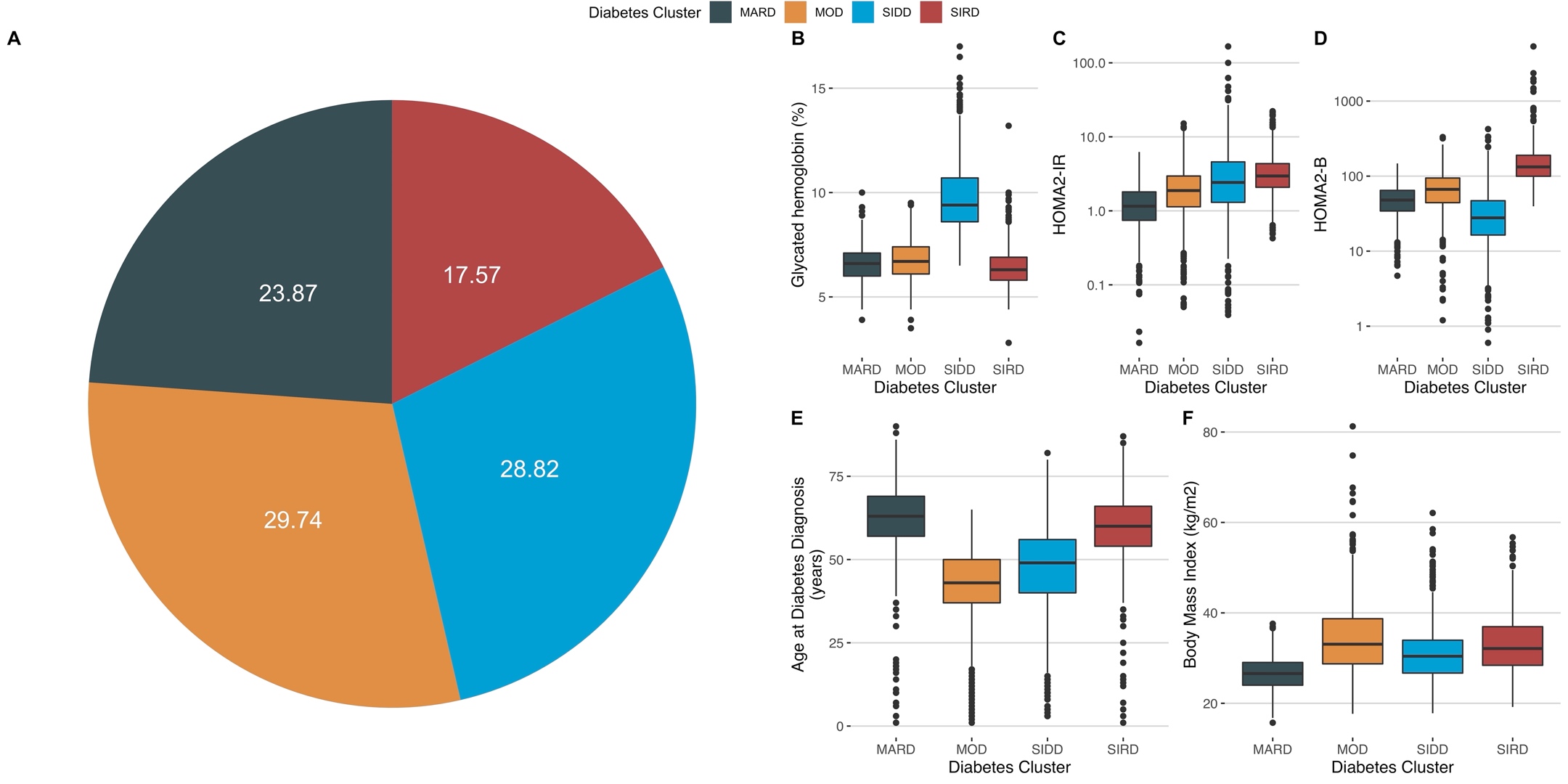

**Supplementary Figure 4:** Prevalence of diabetes subgroups stratified by age categories using National Health and Nutrition Examination Survey data from 1988-2018. *Abbreviations*: MARD, mild age-related diabetes; MOD, mild obesity-related diabetes; SIDD, severe insulin-deficient diabetes; SIRD, severe insulin-resistant diabetes.

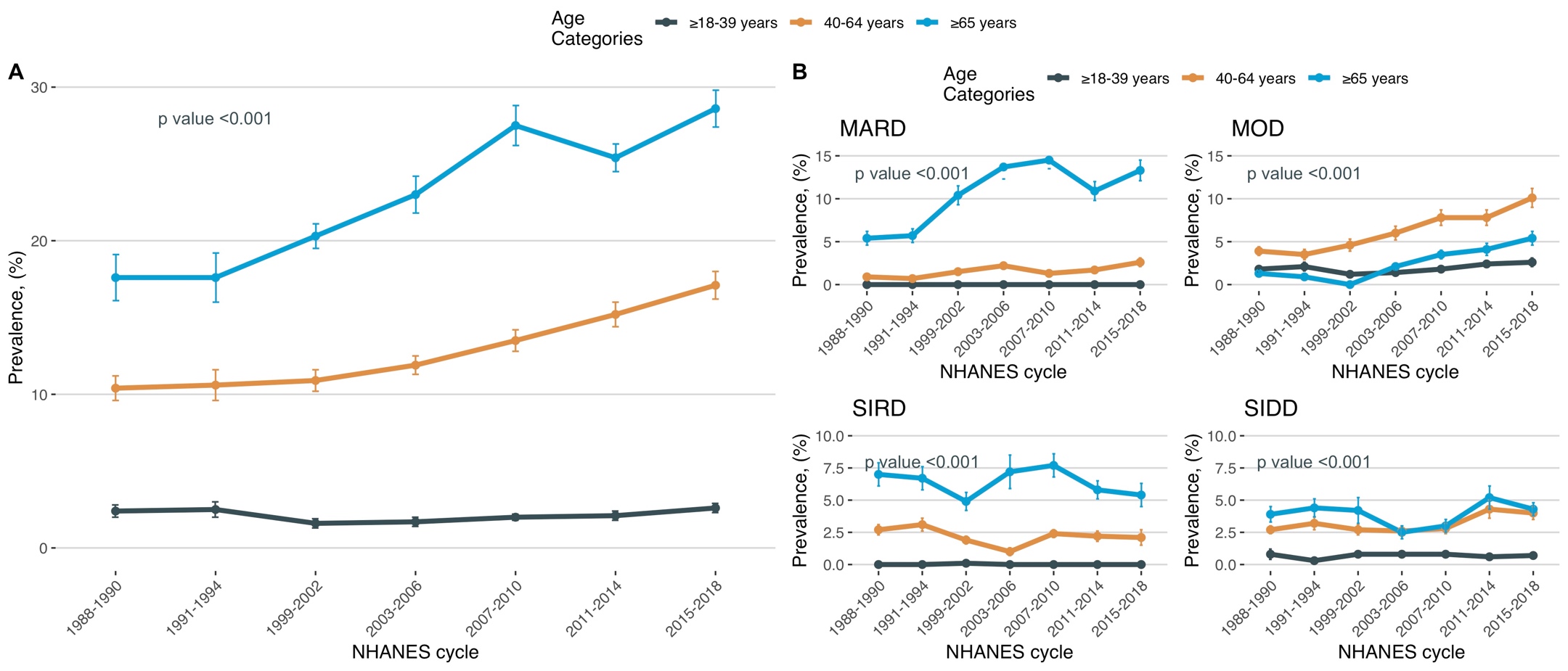

*Footnote: The prevalence was estimated using the examination sample weights from NHANES for participants with available insulin measures according to: Ingram, D. D. et al (2018). National Center for Health Statistics Guidelines for Analysis of Trends. Vital and health statistics. Series 2, Data evaluation and methods research, (179), 1–71*

**Supplementary Figure 5:** Prevalence of diabetes subgroups stratified by BMI categories using National Health and Nutrition Examination Survey data from 1988-2018. *Abbreviations*: MARD, mild age-related diabetes; MOD, mild obesity-related diabetes; SIDD, severe insulin-deficient diabetes; SIRD, severe insulin-resistant diabetes.

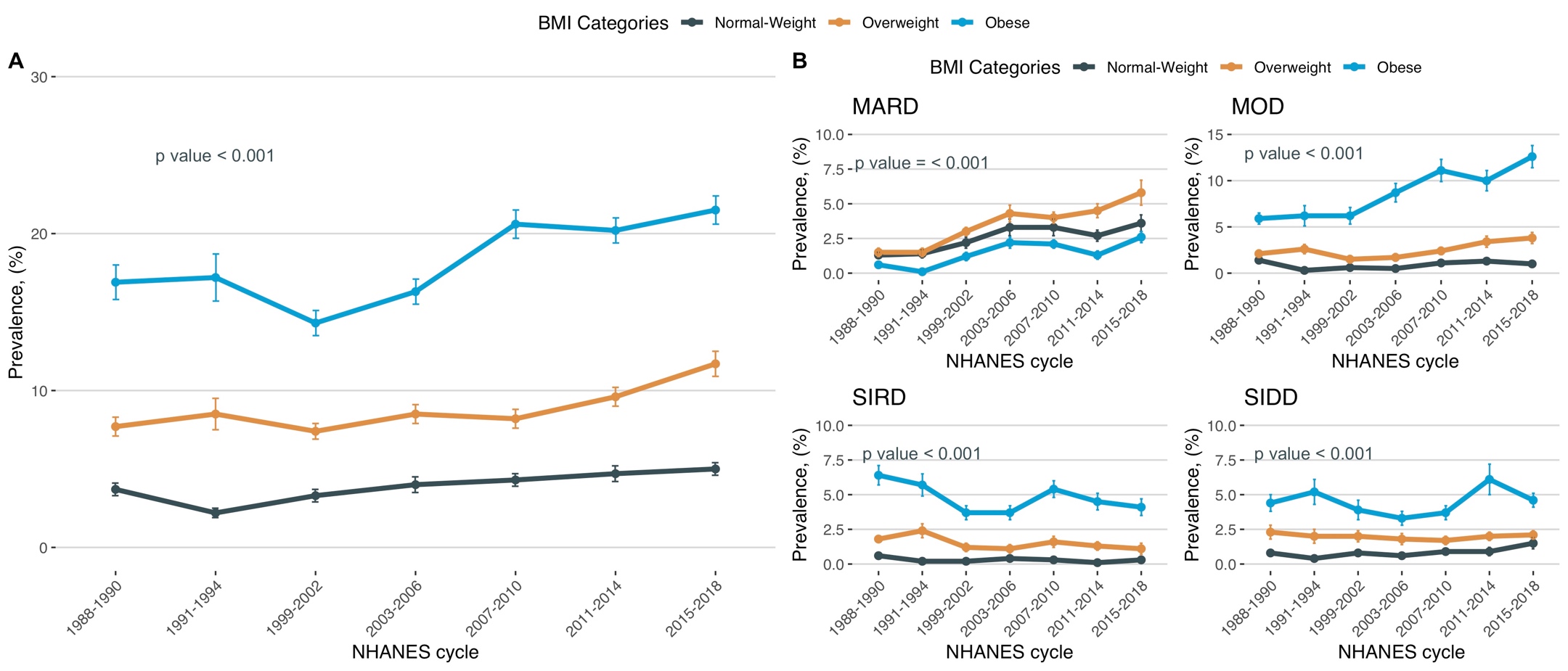

*Footnote: The prevalence was estimated using the examination sample weights from NHANES for participants with available insulin measures according to: Ingram, D. D. et al (2018). National Center for Health Statistics Guidelines for Analysis of Trends. Vital and health statistics. Series 2, Data evaluation and methods research, (179), 1–71*

**Supplementary Figure 6:** Prevalence of diabetes subgroups stratified by time since diabetes onset using National Health and Nutrition Examination Survey data from 1988-2018. *Abbreviations*: MARD, mild age-related diabetes; MOD, mild obesity-related diabetes; SIDD, severe insulin-deficient diabetes; SIRD, severe insulin-resistant diabetes.

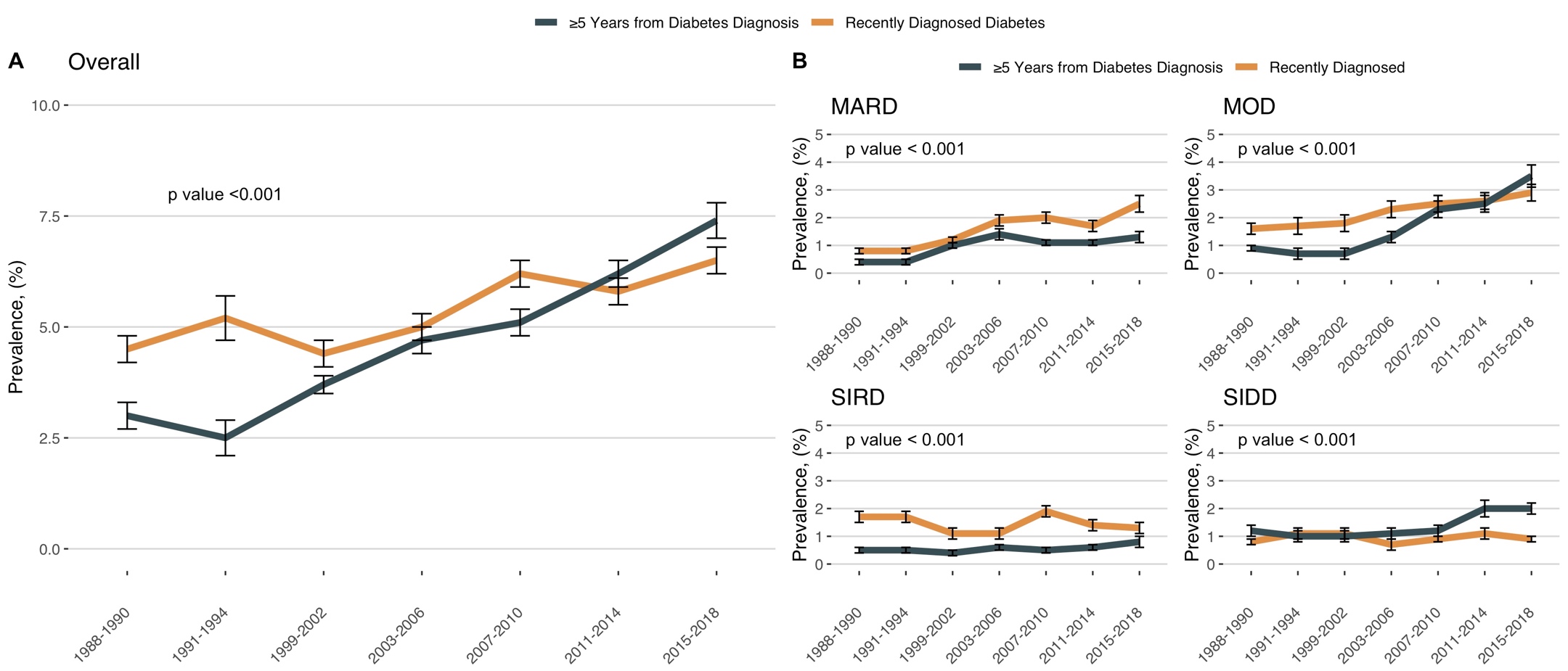

*Footnote: The prevalence was estimated using the examination sample weights from NHANES for participants with available insulin measures according to: Ingram, D. D. et al (2018). National Center for Health Statistics Guidelines for Analysis of Trends. Vital and health statistics. Series 2, Data evaluation and methods research, (179), 1–71*
